# Population health impact, cost-effectiveness, and affordability of community-based HIV treatment and monitoring in South Africa: a health economics modelling study

**DOI:** 10.1101/2022.05.18.22275272

**Authors:** Maitreyi Sahu, Cara J. Bayer, D. Allen Roberts, Heidi van Rooyen, Alastair van Heerden, Maryam Shahmanesh, Stephen Asiimwe, Kombi Sausi, Nsika Sithole, Roger Ying, Darcy W. Rao, Meighan L. Krows, Adrienne E. Shapiro, Jared M. Baeten, Connie Celum, Paul Revill, Ruanne V. Barnabas, for the Delivery Optimization of Antiretroviral Therapy (DO ART) Study Team

**Affiliations:** Department of Health Metrics Sciences, University of Washington, Seattle, WA, USA; Department of Global Health, University of Washington, Seattle, WA, USA; Department of Epidemiology, University of Washington, Seattle, WA, USA; Human Sciences Research Council, Western Cape, South Africa; SAMRC/WITS Developmental Pathways for Health Research Unit, Department of Paediatrics, School of Clinical Medicine, Faculty of Health Sciences, University of the Witwatersrand, Johannesburg, Gauteng, South Africa; Center for Community Based Research, Human Sciences Research Council, KwaZulu-Natal, South Africa; Africa Health Research Institute, KwaZulu-Natal, South Africa; Integrated Community-Based Initiatives, Kabwohe, Uganda; School of Medicine, Yale University, New Haven, CT, USA; Division of Allergy and Infectious Diseases, Department of Medicine, University of Washington, Seattle, WA, USA; Gilead Sciences, Foster City, CA, USA; University of York, York, UK; Division of Infectious Diseases, Massachusetts General Hospital, Boston, MA, USA; Harvard Medical School, Boston, MA, USA

## Abstract

**Introduction:** Community-based delivery and monitoring of antiretroviral therapy (ART) for HIV has the potential to increase viral suppression for individual- and population-level health benefits. However, the cost-effectiveness and budget impact are needed for public health policy.

**Methods and Findings:** We used a mathematical model of HIV transmission in KwaZulu-Natal, South Africa, to estimate population prevalence, incidence, mortality, and disability-adjusted life-years (DALYs) from 2020 to 2060 for two scenarios: 1) standard clinic-based HIV care and 2) five-yearly home testing campaigns with community ART for people not reached by clinic-based care. We parameterised model scenarios using observed community-based ART efficacy. We evaluated incremental cost-effectiveness and net health benefits using a threshold of $750/DALY averted. Sensitivity analyses varied costs of ART, hospitalisation, and testing. Uncertainty ranges (URs) were estimated across 25 best-fitting parameter sets. By 2060, community ART following home testing averted 27.9% (UR: 24.3–31.5) of incident HIV infections, 27.8% (26.8–28.8) of HIV-related deaths, and 18.7% (17.9–19.7) of DALYs compared to standard of care. Adolescent girls and young women aged 15–24 years experienced the greatest reduction in incident HIV (30.7%, 27.1–34.7). In the first five years (2020–2024), community ART required an additional $44.9 million (35.8–50.1) annually, representing 14.3% (11.4–16.0) of the current HIV budget. The cost per DALY averted was $102 (85–117) for community ART compared with standard of care. Providing six-monthly refills instead of quarterly refills further increased cost-effectiveness to $78.5 per DALY averted (62.9–92.8). Cost-effectiveness was robust to sensitivity analyses.

**Conclusions:** In a high-prevalence setting, scale-up of decentralised ART dispensing and monitoring for people not already virally suppressed can provide large population health benefits and is cost-effective in preventing death and disability due to HIV.

## Introduction

Across sub-Saharan Africa, HIV remains a leading cause of death despite universal access to antiretroviral therapy (ART),(1) which substantially reduces HIV-related mortality and transmission and is provided at low cost or free to clients at government clinics.(2,3) However, ART scale-up has fallen short of the 2020 UNAIDS’ target to end the AIDS epidemic, which comprises: 1) identifying 90% of persons with HIV through testing, 2) initiating 90% of people with HIV on ART, and 3) achieving viral suppression among 90% of people starting ART. As of 2021, progress towards global targets stands at 81%, 87%, and 90%, respectively.(4) Men are less likely than women to engage in clinic-based care due to barriers including long wait times, transport costs, and opportunity costs due to missed wages.(5) Untreated HIV and viremia among men is an important driver of the high HIV incidence among adolescent girls and young women, who account for almost one-third (30%) of new infections in eastern and southern Africa despite comprising only one-tenth of the population.(4)

To overcome barriers to ART uptake and adherence, the World Health Organization recommends differentiated service delivery (DSD) models that adapt ART services to patient needs, such as flexible service locations, reduced frequency of refills, and task shifting.(1) Traditionally, these DSD models are recommended only for patients who are already virally suppressed. The recent Delivery Optimization of Antiretroviral Therapy (DO ART) Study evaluated community-based delivery of ART among people who are not already virally suppressed using mobile vans for initiation, monitoring, and resupply in South Africa and Uganda.(6) Community delivery increased viral suppression among men from 54% to 73% – eliminating the gender gap in viral suppression between men and women. The projected cost of community delivery of ART, when provided at scale, was comparable to estimates from clinic-based services. In the context of the COVID-19 pandemic, DSD models including community-based ART with multi-month dispensing were used widely to prevent treatment interruptions.(7)

Data are limited on the population impact, affordability and cost-effectiveness of decentralised ART services when provided at scale and when extended to clients who are not already virally suppressed.(8) Most prior studies of DSD models have estimated either costs or outcomes, but not cost-effectiveness.(9) Two published studies evaluating cost-effectiveness of mobile ART services had mixed findings on cost-effectiveness, but both used higher annual per-patient costs than those for typical modern regimens from the universal test and treat era.(10,11) The objective of this study was to leverage recent outcomes from the DO ART study to parameterise a mathematical model and project population health impact, budget impact, and cost-effectiveness of scaling up community delivery of ART in KwaZulu-Natal, South Africa. This study fills a key gap in the health economics literature on DSD models and community-based ART in particular.(12)

## Methods

### Study design, setting, and outcomes

The DO ART Study was a household-randomised trial conducted in South Africa and Uganda from 2016 to 2019 that evaluated community-based delivery of ART compared with clinic-based ART.(6) Briefly, eligible participants were identified through community-based HIV testing and included adults aged 18 or older living with HIV who were not already on ART or virally suppressed but were clinically stable. Community-based ART consisted of home or mobile van ART initiation with quarterly monitoring and ART refills via mobile vans. The clinic ART group received referral to clinic-based ART initiation and refills. For all groups, supervised lay counsellors conducted HIV testing and counselling and trained nurses provided ART services.

This analysis uses results from the DO ART Study sites in KwaZulu-Natal, South Africa. In KwaZulu-Natal, HIV prevalence is high (27% among adults),(13) 86% of people know their status,(13) and 54% of persons with HIV are virally suppressed.(6) The DO ART Study included 16 communities in the uMgungundlovu and Umkhanyakude Districts of the province of KwaZulu-Natal. Trial results demonstrated that community ART statistically significantly increased viral suppression overall and among men, leading to overall achievement of the 90-90-90 targets; in South Africa, men achieved 72% viral suppression, compared with 51% for the standard of care. A costing analysis embedded in the trial found that community ART, when provided at scale, would cost an estimated $308–312 in the first year and $244–246 in subsequent years, compared with published estimates of $249 for the standard of care in South Africa.(6,14)

### Mathematical model of population health impact

We parameterised an existing deterministic, compartmental model of HIV transmission and disease progression in KwaZulu-Natal with observed viral suppression data from the South African sites in the DO ART Study (Table 1). The DRIVE (Data-driven Recommendations for Interventions against Viral InfEction) model projects the population-level impact of HIV interventions on HIV incidence, prevalence, and mortality. It represents the full KwaZulu-Natal population aged 15–79 years, stratified by age, gender, and ART status. Disease progression is based on CD4 count and viral load. The model captures historical ART rollout and assumes that virally suppressed persons have zero probability of transmitting HIV.(15) The model does not explicitly model ART adherence and discontinuation, but it represents the effect of ART only among individuals who achieve full viral suppression. In addition, the model tracks voluntary male medical circumcision (VMMC) and condom use. Further model details are described in previous publications and in the Supplement Sections I-IV.(16,17)

**Table 1.** Modelled HIV testing and treatment intervention scenarios.

|  | Scenario name | Scenario description | Gender | Persons with HIV who know status (%) | % of persons with HIV who know status who are on ART and virally suppressed | Distribution of people on ART (%)<sup>«</sup> |  | Population-level viral suppression among persons with HIV (%)<sup>¶</sup> |
| --- | --- | --- | --- | --- | --- | --- | --- | --- |
|  |  |  |  |  |  | Clinic ART | Community ART |  |
| 1 | Standard of Care | Clinic ART only | Women | 88.9%<br>[HRSC,(13)] | 70%<br>[DO ART,(6)] | 100% | 0% | 62.2%<br>(88.9% x 70%) |
|  |  |  | Men | 78%<br>[HRSC,(13)] | 51%<br>[DO ART,(6)] | 100% | 0% | 39.8%<br>(78% x 51%) |
| 2 | Home Testing + Community ART | Home HIV testing and counselling campaign once every 5 years + Clinic ART + Community ART for people not reached by clinic care | Women | 93%<sup>†</sup> | 73%<br>[DO ART,(6)] | 91.8%* | 8.2%* | 68%*<br>(93%* x 73%) |
|  |  |  | Men | 92%<sup>†</sup> | 72%<br>[DO ART,(6)] | 68.2%* | 31.8%* | 66%*<br>(92%* x 72%) |

Notes:
\*For Scenario 2, percentages are averages over 2020–2060; because the exact percentage of persons with HIV who know their status fluctuates over time, the distribution of people on ART and population-level viral suppression fluctuate accordingly.
<sup>†</sup>Calculated as baseline HIV testing coverage under Standard of Care, plus additional coverage achieved by home testing and counseling campaigns for persons with HIV not reached by baseline testing. See Supplement, Table S17 for calculations.
<sup>«</sup>See Supplement Section V for calculations.
<sup>¶</sup>Population viral suppression is calculated as: % of persons with HIV diagnosed times % on ART and virally suppressed.

Where possible, model parameters were derived from local empirical data (Supplement Section II). For other parameters, we used approximate Bayesian computation-sequential Monte Carlo (ABC-SMC) to identify parameter sets that best fit observed, historical population-level HIV prevalence (Supplement Section III.a). Projections from the calibrated model are consistent with recent HIV prevalence and HIV incidence estimates, which were not used in calibration (Supplement III.b). Model uncertainty was captured using the range of outcomes generated from the 25 best-fitting parameter sets (referred to as the uncertainty range [UR]). *Model scenarios*

Model scenario assumptions are described in Table 1. We modelled two main scenarios of programme implementation between 2020 and 2060:

1. Baseline HIV testing and quarterly clinic-based HIV care only (“Standard of Care”)

2. Community-based home HIV testing and counselling (HTC) campaigns, conducted once every five years, followed by quarterly community-based ART delivery and monitoring for people who are not reached by the standard of care (“Home Testing + Community ART”)

We modelled viral suppression among persons with HIV on ART for each scenario; details are included in Table 1 and Supplement Section V. For Scenario 2, we assumed that 75% of adults aged 15–79 were reached by five-yearly home testing campaigns as observed in prior studies.(18) For both scenarios, we assumed that 2020 VMMC coverage levels continued through 2060.

In sensitivity analyses, we evaluated two additional scenarios: (1) to determine the impact of home testing alone, we modelled a scenario with home testing and clinic ART; and (2) we explored the impact of increased VMMC on study results (Supplementary Section VII).

### Cost and budget impact

Cost parameters were obtained from the DO ART Study and the literature and inflated to 2020 USD (Table 2). For clinic-based care, we used published cost estimates from South Africa.(14) For community-based ART, we used cost projections from the DO ART Study under a scaled-up scenario.^6^ We also included estimated hospitalisation costs, which varied by CD4 cell count category (Table 2). The cost of HIV testing campaigns was applied to the HIV-negative and undiagnosed HIV-positive population, assuming 75% coverage.(18) To assess affordability of the incremental programme cost of Home Testing + Community ART, we compared the five-year annual incremental undiscounted programme costs to the reported annual programme costs for KwaZulu-Natal in South Africa’s Consolidated Spending Report.(19)

**Table 2:** Primary cost parameters included in the primary and sensitivity analyses.

|  | Parameter | Value | Lower | Upper | Source<br>(Main) | Source<br>(Lower) | Source<br>(Upper) |
| --- | --- | --- | --- | --- | --- | --- | --- |
| 1 | *Home testing for HIV* |  |  |  |  |  |  |
|  | Positive test | 5.81 | 2 | 20.5 | Meyer-Rath,<br>2019<br>(14) | OraQuick self-<br>test<br>(20) | Chang,<br>2016 (SEARCH)<br>(21) |
|  | Negative test | 5.23 | 2 | 20.5 |  |  |  |
| 2 | *Annual HIV-related hospitalisation* |  |  |  |  |  |  |
|  | CD4 <200 | 125.50 | 71.50 | 222.5 | Meyer-Rath,<br>2013<br>(22) | Meyer-Rath,<br>2013<br>(22) | Meyer-Rath,<br>2013<br>(22) |
|  | CD4 200-350 | 58 | 39 | 97 |  |  |  |
|  | CD4 350+ | 39 | 23 | 76 |  |  |  |
|  | On ART | 45 | 23 | 80 |  |  |  |
| 3 | *Community ART* |  |  |  |  |  |  |
|  | First year | 310.84 | 250 | 618.6 | DO ART<br>(at-scale)<br>(6) | DO ART 6-month refill<br>(calculated) | DO ART (as<br>observed)<br>(6) |
|  | Subsequent years | 245.15 | 199 | 500.88 |  |  |  |
| 4 | *Clinic ART* | 249.15 | 109.55 | 315.39 | Meyer-Rath,<br>2019<br>(14) | Nichols, 2021<br>(SDC6)<br>(23) | Thomas, 2021<br>(PopART)<br>(24) |

Notes:
*In the analysis, all costs are inflation-adjusted to 2020 USD. Here they are presented in nominal USD for ease of reference with the literature.*

### Cost-effectiveness and net health benefits

The primary incremental cost-effectiveness ratio (ICER) of interest was the incremental cost per disability-adjusted life-year (DALY) averted. DALYs were calculated cumulatively as the sum of years of life lost (due to all causes) and years of life lived with disability, assuming an average life expectancy of 80 years, health effects truncated at the end of the time horizon in 2060, and disability weights from the 2019 Global Burden of Disease (details in Supplement Section X).(25) For a secondary outcome, we estimated incremental cost per HIV infection averted and per HIV-associated death averted. To compare the health benefits of the intervention with health benefits foregone from not funding other health programmes, we estimated net health benefits (NHB), which is a measure that compares the health gains from committing scarce funding to the intervention to the health opportunity costs of foregone health gains due to the committed resources consequentially being unavailable to deliver other forms of health care. A positive net health benefit indicates that overall population health increases as a result of the programme.(26) We measured NHB at the population level using the following calculation, in which the cost-effectiveness threshold represents the ICER of foregone interventions:

NHB = incremental DALYs averted – (incremental costs / cost-effectiveness threshold) We used a time horizon through 2060 and a cost-effectiveness threshold of $750 as described in South Africa’s HIV investment case.(27) All costs and health outcomes were discounted 3% per year for the cost-effectiveness analysis. For interpretability, we also present undiscounted costs and health outcomes. The uncertainty from the mathematical model was carried through the health economic analysis by performing each calculation across all 25 best fitting sets. Our reporting followed the guidelines set by the Consolidated Health Economic Evaluation Reporting Standards (CHEERS).(28)

### Sensitivity analysis

We conducted one-way sensitivity analyses varying the community-based ART costs, clinic ART costs, hospitalisation costs, and testing costs. A two-way sensitivity analysis included the upper bound cost of community-based ART and lower bound for clinic ART. We also assessed sensitivity to the analytic time horizon and discount rate.

### Software

The mathematical model was implemented in Matlab Version 2020b. The health economic analysis used R version 4.0.2.

### Ethics

Ethical approval for the DO ART Study was granted by the review committees at the Human Sciences Research Council in South Africa and the University of Washington. Participants enrolled in the DO ART Study included adults aged 18 and older who provided written informed consent covering counselling about randomisation, study procedures, and research participant rights.(6) This modelling study used only the aggregate outcomes from the DO ART Study (and not individual participant data), so additional ethical approval was not required.

## Results

### Population health impact

Figure 1 shows the projected cumulative new HIV cases and HIV-associated deaths for the two scenarios, by gender. Five years after programme start, in 2025, Home Testing + Community ART was projected to avert 73,569 HIV-related deaths among men (32.5%, uncertainty range [UR]: 32.2–32.7) and 33,831 HIV-related deaths among women (13.3%, 13.1–13.6); and 32,359 cases among men (17.1%, 16.5–17.8) and 91,362 cases among women (28.0%, 26.6–28.6). By 2060, we estimated that Home Testing + Community ART averted a total of 441,643 HIV-related deaths among men (34.5%, 33.8–35.3) and 432,372 HIV-related deaths among women (23.1%, 22.0–24.3); 322,826 cases among men (25.4%, 22.2–29.0) and 634,982 cases among women (29.4%, 25.4–33.3). Home Testing + Community ART averted 17.5% (14.8–19.0) of DALYs among men and 14.7% (13.9–15.2) among women. In total, we estimated that Home Testing + Community ART averted 13.0 million DALYs compared with the Standard of Care scenario (18.7%, 17.9–19.7). The largest HIV incidence reduction occurred among adolescent girls and young women aged 15–24, among whom we estimated that Community ART averted 357,480 new cases of HIV (30.7%, 27.1–34.7) by 2060 (Supplement Section VI). Among adolescent girls and young women aged 15–24, HIV incidence declined from an estimated 4.2% (2.7–4.9) in 2020 to 2.6% (1.8–3.4) under the Standard of Care and to 1.6% (0.9–2.1) under Home Testing + Community ART in 2060.

**Figure 1:**
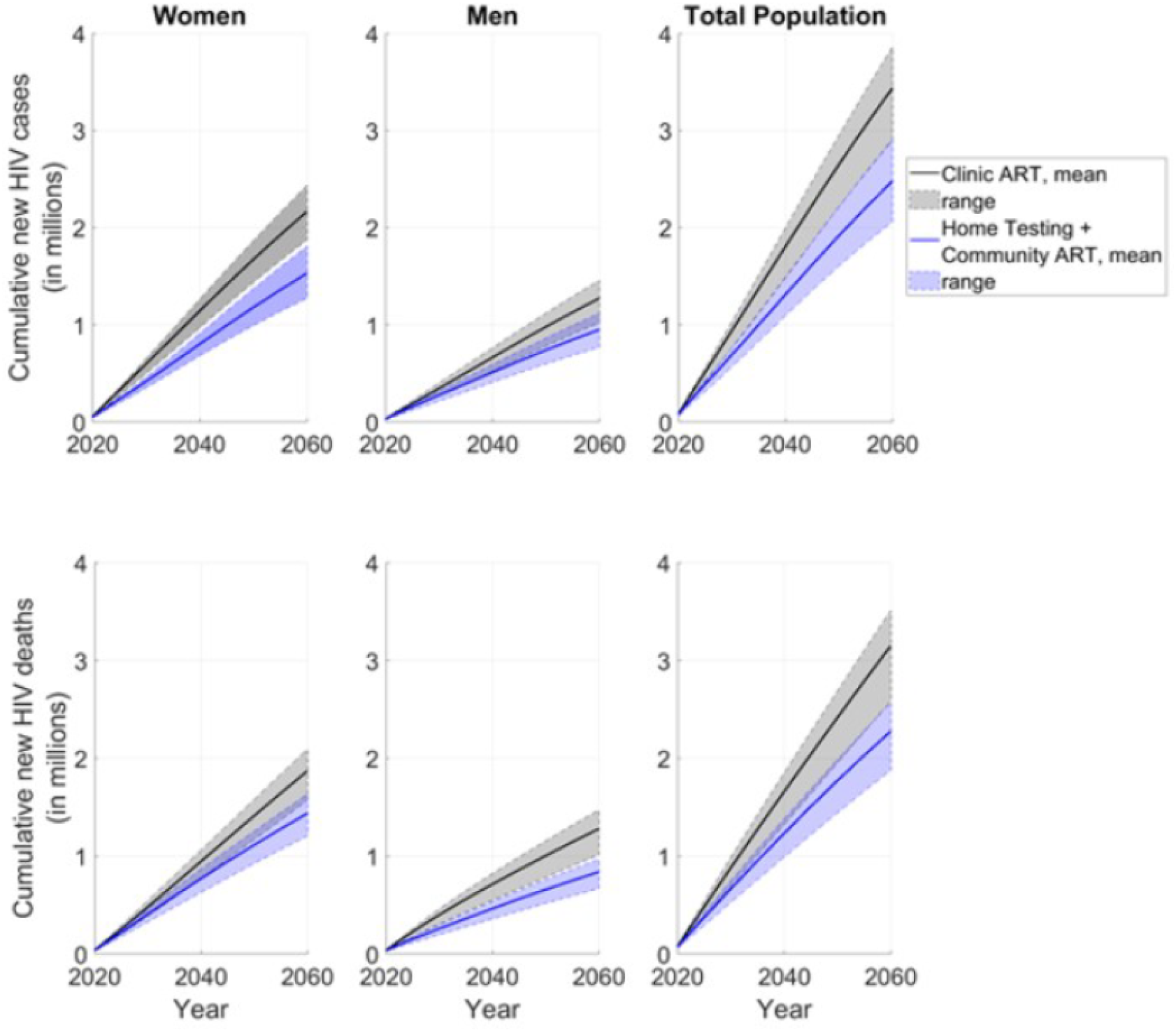
Projected cumulative HIV cases and deaths over time by gender for Standard of Care compared to Home Testing + Community ART. Notes: Error bars for uncertainty ranges represent the minimum and maximum for the 25 best-fitting parameter sets.

### Budget impact

The undiscounted incremental annual programme cost for Home Testing + Community ART for KwaZulu-Natal compared with Standard of Care was estimated to be $31.6 million (UR: 21.8– 40.5) per year (Table 3). During the first five years of the programme (2020–2024), annual programme costs were $44.9 million (35.8–50.1) higher than Standard of Care. When compared with the annual provincial budget for HIV programming, as reported by the Department of Health, of $313.2 million per year, this incremental cost amounted to a 14.3% (11.4–16.0) additional initial investment. For both Standard of Care and Home Testing + Community ART, programme costs for ART delivery were projected to decline between 2020 and 2060 due to reduced prevalence; however, the estimated decline was marginally greater for Home Testing + Community ART (12.7%, 0.6–25.1) compared with Standard of Care (12.5%, -41.0 to 23.5) (Figure 2). By 2049, 56% of model projections estimated lower annual costs for Home Testing + Community ART compared with the Standard of Care; in 2049, costs for Home Testing + Community ART were projected to be $797,261 lower (–15.4 million to 14.2 million) compared to Standard of Care.

**Table 3:** Incremental costs, health outcomes, and cost-effectiveness ICER per infection for HTC + Community ART, compared with Standard of Care, by 2060 (in 2020 USD).

|  | **Home Testing + Community ART, vs Standard of Care** | **Home Testing + Clinic Care only, vs Standard of Care** |
| --- | --- | --- |
| **Cost and budget impact** (*undiscounted*) |  |  |
| Incremental annual programme cost, 2020-2060 | 31.6 million (UR: 21.8–40.5) | 19.5 million (UR: 16.7–22.2) |
| Incremental annual programme cost, 2020-2024 | 44.9 million (UR: 35.8–50.1) | 20.6 million (UR: 17.13–22.59) |
| Initial investment required, 2020-2024 | 14.3% (UR: 11.4–16.0) | 6.4% (UR: 5.3–7.1) |
| **Health gains** (*undiscounted*) |  |  |
| HIV cases averted | 957,808 (UR: 775,441–1,068,738) | 241,950 (UR: 188,780–278,365) |
| HIV deaths averted | 874,015 (UR: 703,693–965,636) | 246,258 (UR: 192,549–275,694) |
| DALYs averted | 13.0 million (UR: 10.5–14.5) | 3.7 million (UR: 2.9–4.2) |
| **Cost-effectiveness** (*discounted 3% for both costs and health gains*) |  |  |
| Cost per case averted | \$1570 (UR: 1206–1950) | \$3346 (UR: 2892–3838) |
| Cost per death averted | \$1748 (UR: 1451–2013) | \$3377 (UR: 3123–3730) |
| Cost per DALY averted | \$102 (UR: 85–117) | \$192 (UR: 181–211) |

Notes:
*Costs are presented in 2020 USD. Values are the mean of 25 best-fitting parameter sets. Uncertainty ranges (URs) represent the minimum and maximum for the 25 best-fitting parameter sets from the model projections.*

**Figure 2:**
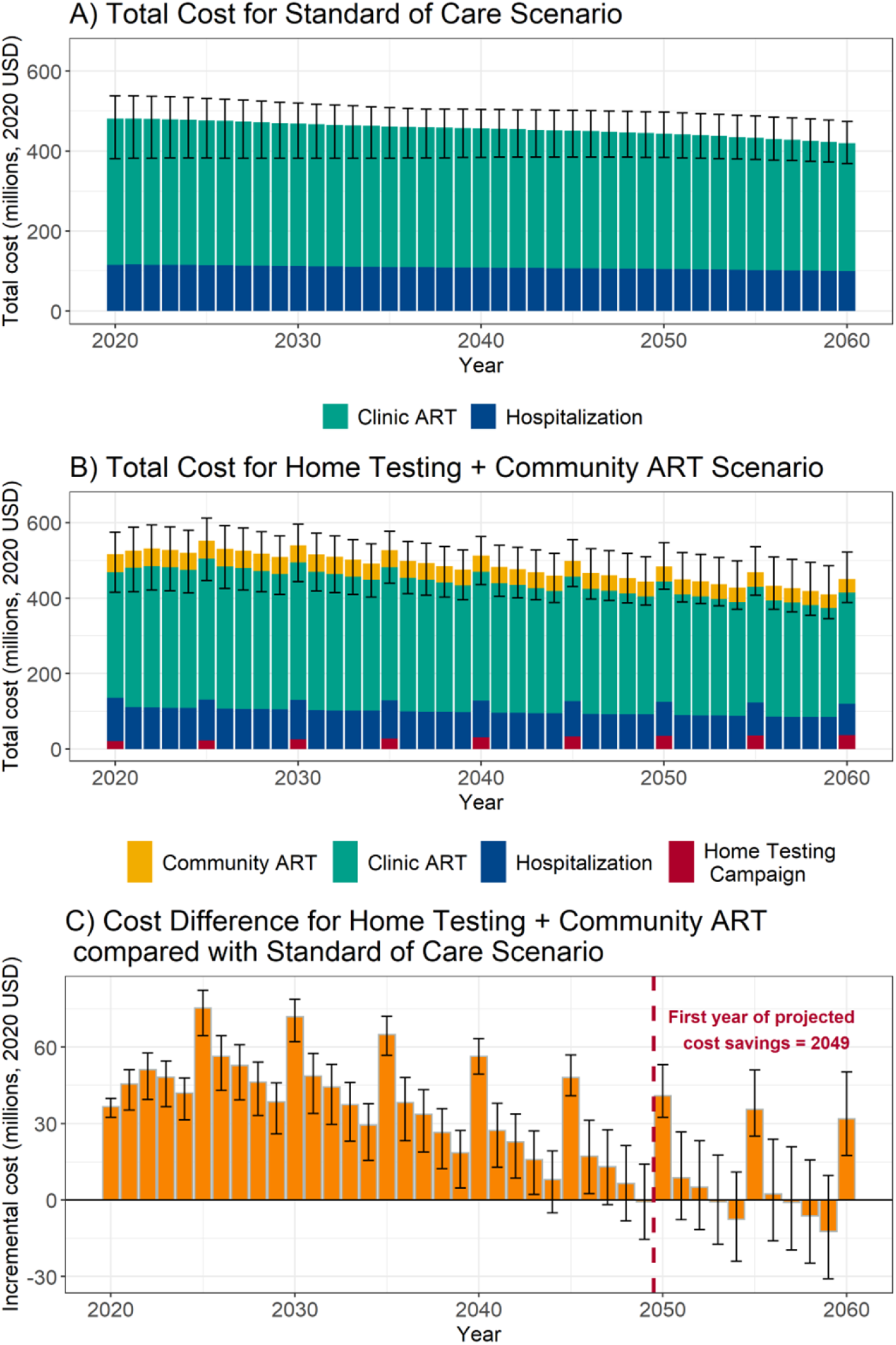
Total and incremental annual ART programme costs for Home Testing + Community ART versus Standard of Care for KwaZulu-Natal province, 2020–2060. Notes: Costs are undiscounted and include ART and downstream hospitalisation costs, but not upstream prevention and baseline testing costs. Values are the mean of 25 best-fitting parameter sets. Error bars for uncertainty ranges represent the minimum and maximum for the 25 best-fitting parameter sets for totals (Panels A and B) and the difference (Panel C).

### Cost-effectiveness and net health benefits

Compared with the Standard of Care, the estimated total incremental cost per DALY averted for Home Testing + Community ART was $102 (UR: 85–117), below the threshold of $750 per DALY averted (shown in Table 3 and Figure 3a). The estimated cost per case averted was $1570 (UR: 1206–1950) and the cost per HIV-associated death averted was $1748 (1451–2013). The net health benefit, measured in terms of DALYs and using a cost-effectiveness threshold of $750, was positive within the first three years of implementation, indicating that the health gains accruing even within three years of implementing community ART would justify the additional costs of delivery (Figure 3b).

**Figure 3:**
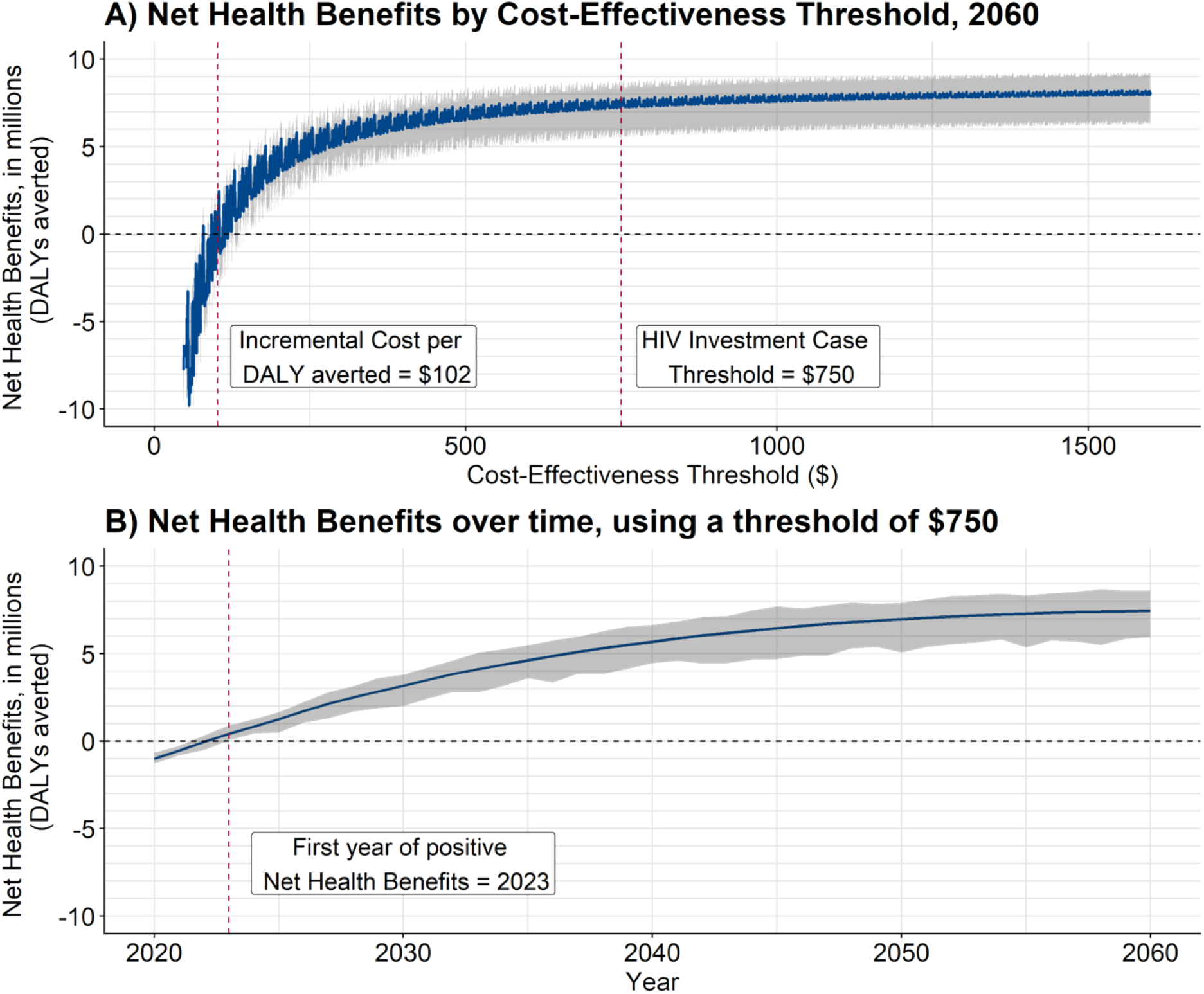
Net health benefits (NHB) of Home Testing + Community ART compared to Standard of Care, by threshold. Notes: A positive NHB implies that overall population health would be increased as a result of the new intervention, while a negative NHB implies that the health benefits of the new intervention are not sufficient to outweigh the health losses from not funding other health programmes in order to fund this intervention. NHB is calculated as the [cumulative incremental DALYs averted – (cumulative incremental costs / cost-effectiveness threshold)]. The discounting rate for costs and health outcomes is 3%. Uncertainty ranges are the minimum and maximum values for the 25 best-fitting model sets.

To separate the impact of Home Testing versus that of Community ART, we also considered a scenario with just Home Testing + Clinic ART. Table 3 shows that home testing alone averted a substantial number of HIV cases (241,950) and deaths (246,258), though these numbers amounted to less than a third of HIV cases and deaths averted by Home Testing + Community ART by 2060 (957,808 and 874,015, respectively).

### Sensitivity analysis

Across all sensitivity analyses, the incremental cost-effectiveness did not cross the threshold of $750 per DALY averted (Figure 4). In the one-way sensitivity analysis of cost parameters, the largest driver of cost-effectiveness was the cost of community ART; using the upper bound for the cost of community ART led to an ICER of $228.2 per DALY averted (UR: 207.6–247.0). Additionally, in a two-way sensitivity analysis using the upper bound cost for community ART and the lower bound cost for clinic ART, the cost per DALY averted was $234.0 (221.1–247.7), below the threshold of $750. In the baseline analysis, community-based ART was cost-effective up to an annual cost of $1616, using a threshold of $750. For the Home Testing + Community ART scenario, increasing the proportion of the population on community-based ART in Scenario 2 (to 100%) but achieving the same health gains led to much greater cost-effectiveness approaching cost saving because the price of community-based ART was cheaper than standard of care after the first year (ICER = $7.9, –4.7 to 19.1). Reducing the time horizon also did not cause the ICER to cross the cost-effectiveness threshold: using a horizon through 2045 and 2030 yielded ICERs of $171 (UR: 157–180) and $400 (UR: 392–413), respectively (Supplement Section XI). A single two-way sensitivity analysis crossed the threshold of $750 per DALY averted: when applying the cost of community-based ART in Scenario 2 to 100% of people on ART and also using the observed DO ART community-based ART costs, the ICER was $812.0 (779.9–856.6); however, this analysis assumed no additional cost efficiencies from applying community-based ART to 100% of the population [Appendix Figure S27]. Incremental cost-effectiveness was not impacted when the level of VMMC scale-up was increased for both scenarios (Supplement Section VII). In a sensitivity analysis which included the cost of six-monthly refills instead of quarterly refills, cost-effectiveness further increased to an ICER of $78.5 (62.9–92.8) per DALY averted.

**Figure 4:**
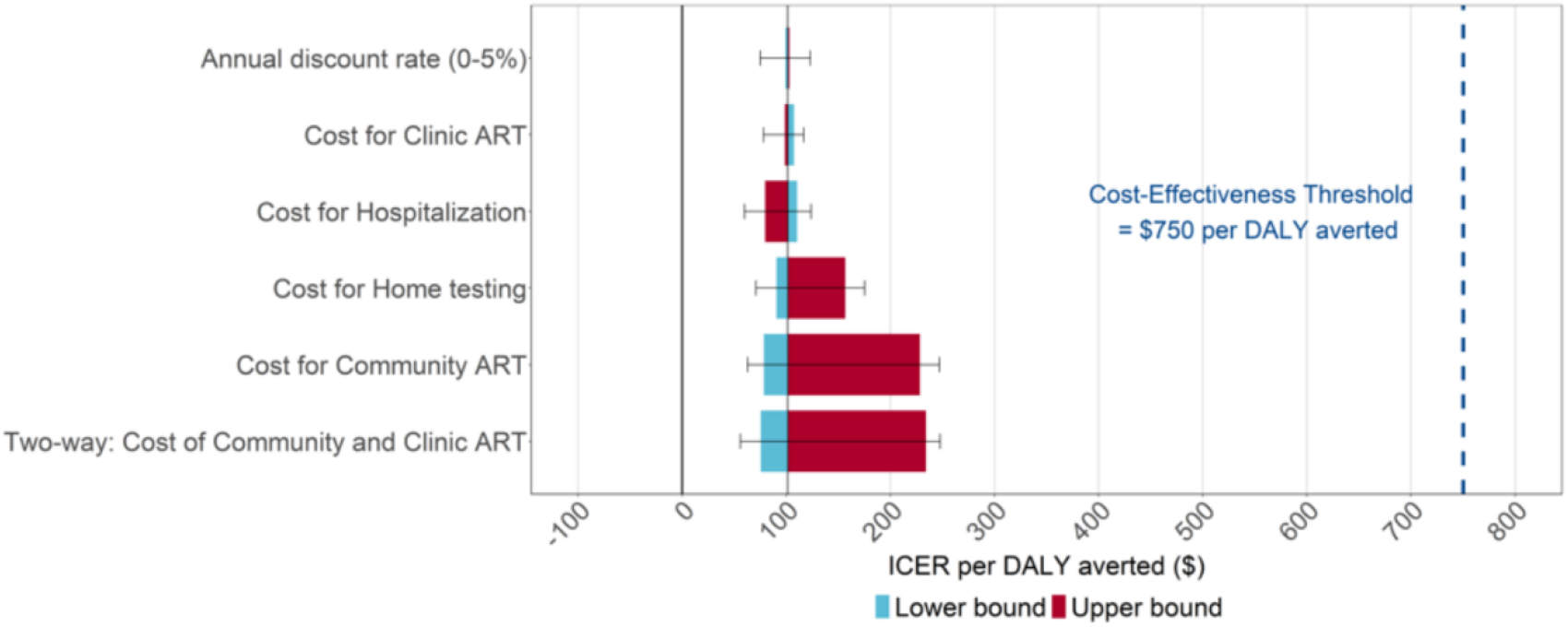
Sensitivity analysis for cost per disability-adjusted life-year (DALY) averted. Notes: Uncertainty ranges are the minimum and maximum values for 25 best-fitting model sets. Incremental cost-effectiveness ratios (ICERs) are presented in 2020 USD. The two-way sensitivity analysis uses the upper bound cost for Community ART and lower bound cost for Clinic ART.

## Discussion

The results from this health economics modelling study provide evidence that community-based delivery of HIV care is cost-effective in high-prevalence settings, with large gains in population health. Specifically, scale-up of Home Testing + Community ART requires an initial investment representing 14.3% of the current annual HIV budget and has potential to avert 28% of HIV-associated deaths, and 28% of HIV infections by 2060, resulting in an ICER of $102 per DALY averted. In addition, within three years of programme implementation the health gains from this programme would justify the additional costs of delivery. Beyond the first few years of implementation and through 2060, adverse health outcomes (incidence, mortality, and DALYs) continue to decline substantially and ART costs decline because of the reduced secondary transmission. Our health economics results are robust across sensitivity analyses, and community-based ART with six-monthly (instead of quarterly) refills can even further increase cost-effectiveness to $78.5 per DALY averted. The benefits of this strategy may be amplified during the COVID-19 pandemic since community-based care allows clients to avoid visiting clinics, decreasing exposure to respiratory illness.

Our modelling study finds remarkable effects on transmission to women. In particular, by 2060 we estimate a decrease of 31% in incident cases among adolescent girls and young women aged 15–24 years, who continue to experience the highest risk of HIV infection compared with other demographic groups.(29) These results are driven by impact of community-based ART on engaging men in care. The clinical results from the DO ART Study demonstrated community delivery of ART substantially increased viral suppression among men from 54% to 73% because men experienced gains from overcoming logistical barriers to facility-based care including lost wages, fixed operating hours, stigma, and gender norms.(5,6) As a result, since virally suppressed men do not transmit HIV, our population modelling demonstrates that the greatest health impact of increased ART uptake among men occurs among adolescent girls and young women – a large driver of the overall cost-effectiveness. Community-based ART has potential to further slow the rate of new infections among adolescent girls and young women alongside ongoing HIV testing, treatment and VMMC programs.(29)

Several recent trials and modelling studies have evaluated effectiveness and cost-effectiveness of community-based HIV testing and linkage to care. A prior modelling study suggested that home testing and counselling with linkage to care reduced incidence by 40.6% in ten years and the incremental cost-effectiveness was $900 per quality-adjusted life-year (QALY) gained.(16) The recent PopART study found that three annual rounds of universal test and treat via home-based testing and counselling cost $326 (266–391) per DALY averted, while 17 annual rounds from 2014 to 2030 cost $625 (538–757) per DALY averted.(24) However, home testing does not guarantee uptake of ART – in a large cohort in South Africa, poor linkages to care persisted following home testing (54% were not verified as linked to care).(30) Same-day community-based initiation of ART has shown promise in improving linkage to care at three months (69% versus 43%) and viral suppression at 12 months (50% versus 34%).(31) The DO ART Study, which evaluated community-based ART initiation and resupply, found even further gains in viral suppression at 12 months for men (73% versus 54%). Thus, this study’s findings that community-based initiation and resupply, when combined with home testing and counselling, leads to additional large gains in population health impact and cost-effectiveness beyond that of home testing alone, is in line with current literature and findings.(32,33)

This study adds to the evidence for extending DSD to key populations beyond clinically stable clients who already achieved viral suppression. WHO currently recommends various DSD models which are adapted to client needs to improve ART initiation and retention, but these recommendations are not well defined and are generally only provided for clinically stable patients.(1) Recent advocacy suggests that DSD models could achieve even greater outcomes if adapted based on the sub-population (e.g. men, women, adolescents, key populations), context (prevalence level, urban/rural, etc.), and clinical characteristics (co-morbidities, clinically unstable, clinically stable).(34) We find that in a high-prevalence setting, community-based ART is a cost-effective strategy to engage virally unsuppressed men in care and in the long term can improve health outcomes for both men and women.

We acknowledge that this research has limitations. First, we do not evaluate the comparative cost-effectiveness of various DSD models such as community adherence groups and task shifting, or different forms of ART such as long-acting injectables. We also do not model key prevention interventions such as oral and injectable PrEP; however, since the outcome of interest is incremental cost-effectiveness, scale-up of prevention does not affect the health economics results. When we include VMMC in the model (Supplement Section VII), the ICER for community-based ART remains exactly the same because scale-up is the same in both scenarios. Second, community-based ART costs are projected for a scaled-up programme and require client volume to be achievable. However, our results are robust to using the higher cost of community-based ART observed in the DO ART Study. Third, this study is limited to a high-prevalence setting in South Africa; while we expect the results to be generalisable to other high-prevalence settings in sub-Saharan Africa, results may differ in settings with lower HIV prevalence. Finally, we do not specifically model the impact of the COVID-19 pandemic, which could affect both outcomes and costs. Since the analysis is conducted over a long time horizon, we do not expect the pandemic to affect our interpretation.

South Africa has ongoing demonstration projects for community delivery of HIV care, which incorporates several components of DSD. This research supports continued implementation and evaluation of community-based options for ART initiation, monitoring, and resupply, including mobile vans and pickup points (such as smart lockers). These options can be offered at the time of HIV testing/ART initiation so that the service is client-centered; ideally clients would have a choice regarding the mechanism for ART delivery and monitoring. In this way, by leveraging existing high rates of HIV testing and expanding differentiated services to those with detectable viral load, we can increase linkage to care and viral suppression. This modelling analysis supports decentralised ART dispensing and monitoring as a public health approach with broad individual- and population-level benefits, including improved viral suppression among men and reduced transmission to adolescent girls and young women.

## Data Availability

The data used for this study is available upon request.

## Contributors

RVB, MS, CJB, DAR, and PR conceptualised the project and the modelling strategy. CJB, DWR, RY, and RVB developed the mathematical model. MS and DAR conducted health economic analysis. RVB, PR, and DAR supervised the analysis. MS and CJB generated tables and figures. MS wrote the first draft of the manuscript; CJB drafted the technical appendix. All authors revised and approved the manuscript for content.

## Data sharing

The data used for this study is available upon request.

## Acknowledgements

The authors would like to thank the DO ART Study Team and the participants who participated in the trial. We also thank Adrienne Chew for scientific editing support, and Marina Antillon for technical guidance. This work was facilitated though the use of advanced computational, storage, and networking infrastructure provided by the Hyak supercomputer system at the University of Washington.

